# Accelerated partner therapy contact tracing for people with chlamydia: The LUSTRUM cluster cross-over randomised controlled trial

**DOI:** 10.1101/2021.08.04.21261369

**Authors:** Core writing group, Claudia S Estcourt, Andrew Copas, Nicola Low, Fiona Mapp, Oliver Stirrup, the LUSTRUM research programme, Jackie Cassell

## Abstract

**Objective:** To assess accelerated partner therapy (APT) as a contact tracing intervention for people with chlamydia.

**Design:** Cross-over cluster-randomised controlled trial.

**Setting:** 17 sexual health clinics (clusters) in the United Kingdom, 2018-2019.

**Participants:** Heterosexual people aged over 16 years with a positive *Chlamydia trachomatis* test result and/or clinical diagnosis of pelvic inflammatory disease, cervicitis, non-gonococcal urethritis or epididymo-orchitis, and reporting one or more contactable sexual partner in the past six months, and their sexual partners.

**Interventions:** Clusters were assigned by random permutation to either (a) usual care: health care professional advises the index patient to tell their sex partner(s) to attend clinic for sexually transmitted infection (STI) screening and treatment, or (b) usual care plus the offer of APT: healthcare professional assesses sex partner(s) by telephone, then sends or gives the index patient antibiotics and STI self-sampling kits for their sex partner(s). After a two-week washout period, clinics crossed over to the opposite exposure. Each period lasted 6 months.

**Main outcome measures:** The primary outcome was the proportion of index patients with a positive *C. trachomatis* test 12-24 weeks after treatment. Secondary outcomes included proportions and types of sex partners treated.

**Analysis:** Intention-to-treat, fitting random effects logistic regression models.

**Results:** All clinics completed both periods. Overall, 1536 and 1724 recruited index patients provided data in intervention and control phases respectively. In total, 4807 sex partners were reported, of whom 1636 (34%) were committed/established partners. Overall, 293/1536 (19.1%) of index patients chose APT for a total of 305 partners, of whom 248 accepted. In intervention and control phases, 666 (43%) and 800 (46%) of index patients were tested for *C. trachomatis* at 12-24 weeks; 31 (4.7%) and 53 (6.6%) were positive, adjusted odds ratio (aOR) 0.66 (95% CI 0.41–1.04, p=0.07). Among index patients with treatment status recorded, the proportion with ≥1 sex partner treated was 775 (88.0%) in the intervention and 760 (84.6%) in the control phase, aOR 1.27 (95% CI 0.96–1.68, p=0.10). Seven adverse events of low severity were recorded.

**Conclusions:** APT can be safely offered as a contact tracing option for people with *C. trachomatis* and might reduce the risk of repeat infection. Future research should find ways to increase uptake and develop alternative interventions for one-off partners.

**Trial registration:** ISRCTN15996256

**Ethical approval:** London - Chelsea Research Ethics Committee (18/LO/0773)

**Protocol:** doi: 10.1136/bmjopen-2019-034806

**What is already known on this topic:**

- Contact tracing (partner notification) for chlamydia is a key element of infection control in the population but achieving even modest outcomes can be challenging.
- Accelerated partner therapy (APT) is a contact tracing intervention that provides testing and treatment for sex partners without the need for a face-to-face consultation.
- Pilot studies of APT found improvements in patient-reported outcomes of contact tracing but evidence about biological outcomes is required and the types of sexual partnerships benefitting most from APT are unknown.

**What this study adds:**

- The offer of APT as an additional contact tracing method to usual care likely caused a small reduction in repeat chlamydia infection 12-24 weeks after treatment and an increase in proportion of sex partners treated, compared with usual care alone.
- APT can be safely offered as a cost-saving contact tracing option for heterosexual people with chlamydia and might reduce the risk of repeat infection, particularly for those in emotionally connected relationships, although uptake needs to be improved and novel approaches are needed for one-off partners.

## INTRODUCTION

Contact tracing, also referred to as partner notification (PN), is the process of identifying, testing and treating sex partners of a person diagnosed with a sexually transmitted infection (STI).^1^ Contact tracing is a key element of STI control on several levels.^2^ It should benefit the individual diagnosed with the STI (the index patient) by preventing re-infection, the sex partner who might be the source of infection or could transmit undiagnosed infections to new sexual partners, and it should help to reduce spread of STIs in sexual networks and populations.^3^ *Chlamydia trachomatis* (chlamydia), is the most commonly reported bacterial STI in the United Kingdom (UK),^4–6^ with 229,441 diagnosed cases in England alone in 2019^4^ Most infections are asymptomatic and easily treatable with oral antibiotics. However, untreated infections can lead to pelvic inflammatory disease (PID), infertility, ectopic pregnancy and chronic pelvic pain in women, and epididymo-orchitis in men.^7^ Chlamydia infections do not induce lasting immunity after antibiotic treatment, which is a particular challenge for STI control efforts. In prospective studies with active follow up, around 20% of women have a repeat diagnosis of chlamydia infection in the year after treatment,^8–10^ with a peak incidence at 2-5 months.^11^ Mathematical modelling has shown that improving contact tracing outcomes for chlamydia would be more cost-effective than increasing the coverage of chlamydia testing.^2^

However, contact tracing can be challenging both for patients, who may face barriers to informing sex partner(s) about the STI, and for clinicians, who need time to elicit and discuss sensitive information. Healthcare professionals in British sexual health services often struggle to meet modest outcomes.^12–14^ For people with chlamydia, contact tracing is most often performed through enhanced patient referral,^15,16^ in which a healthcare professional advises the person with an STI (the index patient) to inform their sex partner(s) of the need for testing (routinely, a comprehensive STI & HIV screen^17^) and chlamydia treatment and provides printed/website information. Sometimes the health care professional contacts the sex partner directly (provider referral) without disclosing the identity of the index patient. Allowing patients to choose the most acceptable contact tracing method, which might differ for different sex partners, is considered optimal practice^18^ but recent pressures on UK sexual health services have resulted in a deprioritisation of contact tracing and reduced choice.^19^

We previously developed a new intervention, known as accelerated partner therapy (APT), to improve and speed up the contact tracing process.^20–22^ APT is adapted from expedited partner therapy (EPT), an intervention developed in the United States of America (USA) that improves contact tracing outcomes.^12,23,24^ EPT does not meet UK prescribing guidance^25^ because practitioners provide medication or a prescription for sex partners without any consultation with or prior knowledge of the sex partner. We piloted an early form of APT, in which the healthcare professional telephoned the sex partner, in private, during the index patient’s clinic attendance.^20,21^ The index patient was then given a pack containing antibiotics, chlamydia and gonorrhoea self-sampling kits and an invitation to visit a sexual health clinic for syphilis and HIV testing (requiring venepuncture) to deliver to their sex partner. APT resulted in faster sex partner treatment and greater overall numbers of sex partners treated, when compared with usual practice, but levels of testing for HIV and other STIs were low.(ibid) Since these early studies, HIV and syphilis finger prick blood self-sampling kits have been approved.^26^ The Limiting Undetected Sexually Transmitted infections to RedUce Morbidity (LUSTRUM) Trial aimed to investigate the effects of APT *in addition* to usual practice contact tracing, compared with usual practice alone, in heterosexual people with chlamydia, on the proportion of index patients with a positive *C. trachomatis* test result 12-24 weeks after treatment, and the proportion of sex partners treated, overall and according to sex partner type.^27^ An integral process evaluation^28^ and health economics analysis based on a transmission dynamic model are published elsewhere.^29^

## METHODS

Detailed methods for the LUSTRUM APT trial have been published elsewhere.^27^ Briefly, we did an unmasked cluster cross-over randomised controlled trial (RCT) comparing APT, when offered in addition to usual contact tracing (usual care), with usual care alone. We chose this design because individual randomisation carried a high risk of contamination of the intervention and was operationally unfeasible. The cross-over design allowed each clinic to test both the intervention and usual care and provides efficiencies in patient enrolment. Service-level consent aimed to make the intervention easier and more realistic to deliver in busy clinical settings.

### Population

Eligible index patient participants were aged 16 years or older with a positive test for *C. trachomatis* and/or a clinical diagnosis of conditions for which presumptive chlamydia treatment and contact tracing is initially provided: pelvic inflammatory disease (PID) or cervicitis (women), or non-gonococcal urethritis (NGU) or epididymo-orchitis (men) *and* reported at least one contactable sexual partner in the past six months. Index patients whose test results were subsequently negative for *C. trachomatis* were not included in the statistical analysis. We excluded men who have sex with men (as contact tracing needs may be different), and people with complex clinical, social or other circumstances, such as sexual assault, or with insufficient English language skills to safely engage in telephone consultations.

Eligible partners were defined as a sex partner of the index patient aged 16 years or older, within the appropriate lookback period (six months for chlamydia, three months for PID, one month for NGU),^17^ and selected by the index patient for APT.

### Settings

We enrolled 17 NHS (publicly funded, free to access) sexual health clinics (clusters) across England and Scotland. Clinic selection was based on numbers of reported chlamydia diagnoses data in the Public Health England GUMCAD STI Surveillance System^30^ (England) and geographical diversity (Scotland) to ensure representation from each of three groups: London, non-London metropolitan “cities” and non-London urban “towns”.

### Randomisation

We allocated clinics to intervention or control arm through random permutation within strata, using computer-generated random numbers.^31^ Fourteen clinics were initially randomised, including three strata that were pairs of clinics within one NHS trust (hospital group), one stratum containing five clinics from large cities and another containing three clinics from smaller towns. A further pair and then the final clinic (allocated through simple randomisation) were randomised later, to boost recruitment. To remove the potential for allocation bias, one statistician generated the allocation codes and another randomly permuted the clinic names within the strata. Only then, a third person matched the two to reveal the allocations and inform the trial.

There was a four-month rolling clinic set-up (July – October 2018), then nine clinics entered the intervention phase and eight entered the control phase. At the end of the first six-month period (November 2018 – April 2019) clinics followed their usual contact tracing procedures for a two-week washout period. Then, clinics crossed over to the opposite phase for May – November 2019. For the three clinics randomised later (February 2019), trial periods were condensed to complete recruitment in November 2019. The total duration of the trial was 19 months, allowing for a three-month follow-up period to complete data collection.

### Patient enrolment and data management within the clinic setting

During the initial contact tracing consultation with the index patient, a healthcare professional assessed eligibility for the study for all patients with a positive laboratory test result for *C. trachomatis* or a relevant clinical diagnosis. Healthcare professionals were asked to record their consultations in real time. This included a new classification of sex partner types, developed before the trial.^32^ They used RELAY, a secure web-based data collection platform, developed for this trial based on pilot studies.^20,21^ It was hosted on secure servers and complied with NHS data storage requirements. RELAY was also intended for baseline data collection but, at almost all sites, healthcare professionals pre-screened index patients for eligibility and only created a RELAY record if the index patient met eligibility criteria. Several sites restricted enrolment to a small number of clinical sessions per week.

### Intervention

APT is a complex intervention,^33,34^ which was offered as an additional option, alongside usual care, described in BOX 1 for index patients, and BOX 2 for sex partners. The index patient could choose APT or usual care for each partner. Follow-up was the same for both interventions. If APT was not feasible (e.g. the sex partner could not be reached), usual care was offered instead.

During the control phase, clinics followed their standard protocols for usual care, which in all clinics was enhanced patient referral. Follow-up telephone calls and repeat testing were the same as those during the APT phase.

#### BOX 1

**Overview of APT for index patients**

1. Index patient has contact tracing consultation with healthcare professional (HCP); HCP assesses eligibility for APT.
2. Eligible index patient is offered APT alongside the clinic’s other standard contact tracing options. The patient can choose different methods for different partners.
3. Index patient telephones or messages sex partner (with or without the HCP present, according to preference) to offer immediate telephone assessment by the HCP.
4. Index patient waits in clinic while the HCP conducts APT telephone consultation in private with sex partner. If sex partner accepts APT, the HCP gives the index patient an APT pack (Figure 2) to deliver to their partner and demonstrates how it should be used, or sends the pack to the sex partner directly.
5. Index patient is informed that they will receive a follow-up telephone call in two weeks and they will either receive a chlamydia self-sampling postal kit in 12-24 weeks (preferred), or they may re-attend the clinic for testing.
6. Two-week follow-up: Research Health Adviser (RHA) telephones index patient 2-4 weeks after the consultation to find out about contact tracing outcomes with partner(s), to remind them of the repeat test, and to invite them to be contacted about taking part in a telephone interview regarding their experiences of APT (process evaluation).
7. 12 weeks: Index patient is sent a personalised text reminder about repeat test.
8. 13 weeks: Index patient is sent a self-sample kit by The Doctors’ Laboratory (TDL), London, UK. Index patient returns self-collected sample or attends clinic for repeat testing. S/he receives results either via text message from TDL (negative results) or using routine clinic systems (positive or equivocal results). Positive results are managed according to routine clinic protocol. If the index patient does not return a self-sample or attend clinic for repeat testing, they receive a personalised text reminder eight days after the self-sample kit is sent out, followed by a telephone call from the RHA 13 days after the self-sample kit is sent out. Self-samples received >24 weeks post-contact tracing interview are excluded.

#### BOX 2

**Overview of APT for sex partners**

1. Index patient telephones sex partner to inform them about exposure to chlamydia and offer immediate telephone assessment (APT).
2. If sex partner agrees to APT, HCP telephones them and conducts a clinical assessment in private. If appropriate, sex partner is offered an APT pack (delivered by the index patient or mailed directly). Sex partners for whom APT is inappropriate or who do not wish to continue with the APT option, will be advised by the HCP to attend clinic for further management. During the same telephone call, HCP invites sex partner to be contacted about taking part in a telephone interview regarding their experiences of APT (process evaluation).
3. Sex partner receives APT pack (FIGURE X), which contains: antibiotics (either azithromycin or doxycycline, depending on local clinic practice); condoms; information about chlamydia, gonorrhoea, HIV and syphilis; chlamydia and gonorrhoea self-sampling kit (urine/vulvo-vaginal swab), HIV and syphilis self-sampling kit (finger-prick blood sample), and information leaflet about how to take a sample (including link to an explanatory online video: lustrum.org.uk/test-and-treat); request form for the sample to be processed by the lab; envelope for return of self-sampling kits; and APT pack packaging (envelope or small box, no branding or other identifiable markings, and which fits through standard letterbox).
5. Sex partner completes self-sampling, labels, and returns samples for testing.
6. Sex partner takes antibiotic treatment.
7. Sex partner informed of test results by text (negative results) or routine clinic processes; positive results are managed according to routine care.

### Usual care

#### Outcomes

The primary outcome was the proportion of index patients with a positive test result for *C. trachomatis* 12-24 weeks after the contact tracing consultation, as a proxy for re-infection from an untreated partner. The procedure for obtaining samples is described in BOX 1. The key secondary outcome was the proportion of sex partners treated 2-4 weeks after the initial contact tracing consultation. Other secondary outcomes were: one or more partners treated per index patient; time to partner treatment; proportion of partners notified; and one or more partners notified per index patient, ascertained during a telephone call with a research health adviser. The secondary outcomes of ‘numbers of partners treated/notified’ per index patient were interpreted in the statistical analysis plan as ‘one or more partners treated/notified per index patient’ to match UK reporting standards^17^ and make analysis more tractable with respect to missing outcome data at the partner level.

The health economic evaluation included a cost consequence analysis (described in this report, APPENDIX 1) and a model-based cost-effectiveness analysis reported separately.^29^ We also conducted a process evaluation to explain how APT worked in practice and to contextualise trial results^28^ summarised below.

#### Sample size calculation

The sample size calculation was based on enrolling an average of 160 index patients per clinic per trial phase from 17 clinical services (total 5440 patients) and a coefficient of variation in the number enrolled of 0.5. We expected that 10-25% of patients in the control arm would have *C. trachomatis* detected at the 12-24 week follow-up.^12,35^ We expected that 50% of enrolled patients (80 per clinic per phase and 2720 total) would contribute to the analysis of the primary outcome, assuming that 60% provided a repeat sample and excluding those without confirmed *C. trachomatis* at baseline. This sample size would provide 80% power (at the 5% significance level) to detect a fall in *C. trachomatis* positivity from 10% to 5%, and 82% power to detect a fall from 25 to 17% under the intervention. It provides 87% power to detect an increase from 60% in the control arm to 70% in the intervention arm in the patient-reported outcome of proportion of index patients with one or more partner treated.^13^ Sample size calculations were guided by Giraudeau et al.^36^ but performed conservatively as if the trial were a standard cluster RCT with 17 clinics in each arm. Our calculations assumed an intra-cluster correlation (ICC) of 0.02, in the absence of published data.

### Statistical analysis

Statistical analyses followed a plan agreed before the completion of data collection. The primary analysis was by intention-to-treat, including all recorded eligible patients within study periods. For the primary outcome, and other quantitative outcomes, we fitted mixed effects logistic regression models with fixed effect for intervention condition, with random effects to acknowledge the clustering of index patients for each clinic and for each period nested within clinics.^37^ The intervention effect is expressed as an odds ratio (OR) with 95% confidence interval (CI). Models for secondary outcomes with outcomes quantified for each sexual contact included additional random effects for each index patient. The primary outcome measures used the observed data, adjusted for patient characteristics. We conducted multiple imputation of sex partner treatment status and index patient repeat test results under the ‘missing at random’ assumption using information on index patient sex, ethnicity, enrolment based on presence of NGU and age, and carried out further sensitivity analyses in which we will allowed those lost to-follow-up to be more, and then less, likely to be chlamydia positive at repeat test than those not lost to follow-up.^38^ The analysis of the primary outcome and secondary outcome of one or more sex partner treated per index patient were repeated after exclusion of clinics with very low uptake of the APT intervention (defined after data collection as those with proportion of IPs with APT accepted for at least one partner below 15%). Further details are provided in the supplementary appendix.

### Trial registration and approval

The LUSTRUM trial is registered as ISRCTN 15996256.^39^ After the start of the trial the protocol was changed to allow inclusion and testing of index patient STI samples up to 24 weeks after initial contact tracing consultation. This was changed as trial monitoring indicated some index patients returned their self-sampling kits after 16 weeks (the original cut-off) due in part to a computer server error which sent reminder texts to some index patients later than scheduled. Post hoc analyses are stated.

### Patient and public involvement

Contributors from the public assisted with study design, particularly in relation to the decisions to seek service-level consent, the development of a GDPR-compliant research data opt-out process (D. Rosen (PPI group))^40^ and to offer APT as an additional contact tracing option alongside standard care (C. Ward, D. Rosen(PPI group)). Other contributions addressed the development of all study materials for patients, particularly the design of the APT pack, and videos for the public about the trial^33^ (C Ward, (PPI group)), attendance at key programme research meetings (N. Sutherland, D. Crundwell) and review of various trial reporting documentation.

## RESULTS

Over the study period, clinic level administrative data showed that there were 16,445 chlamydia diagnoses made in potentially eligible people. All 17 clinics completed both trial periods and a total of 1724 and 1536 index patients were included in the analysis of the control and intervention phases, respectively (Figure 1). Participants in intervention and control phases were comparable (Table 1). In the control phase, participants reported a total of 2589 eligible sex partners, (median 1, interquartile range, IQR, 1-2, range 1-20, 66% male). Health professionals categorised these partners as: committed/established 880 (34%), new relationship 342 (13%), occasional 687 (27%), or one-off 680 (26%) (Table 2). In the intervention phase, participants described a total of 2218 eligible sex partners (median 1, IQR 1-2, range 1-10, 64% male). These partners were: committed/established 756 (34%), new relationship 343 (15%), occasional 610 (28%), one-off 509 (23%) (Table 2).

**Figure 1:**
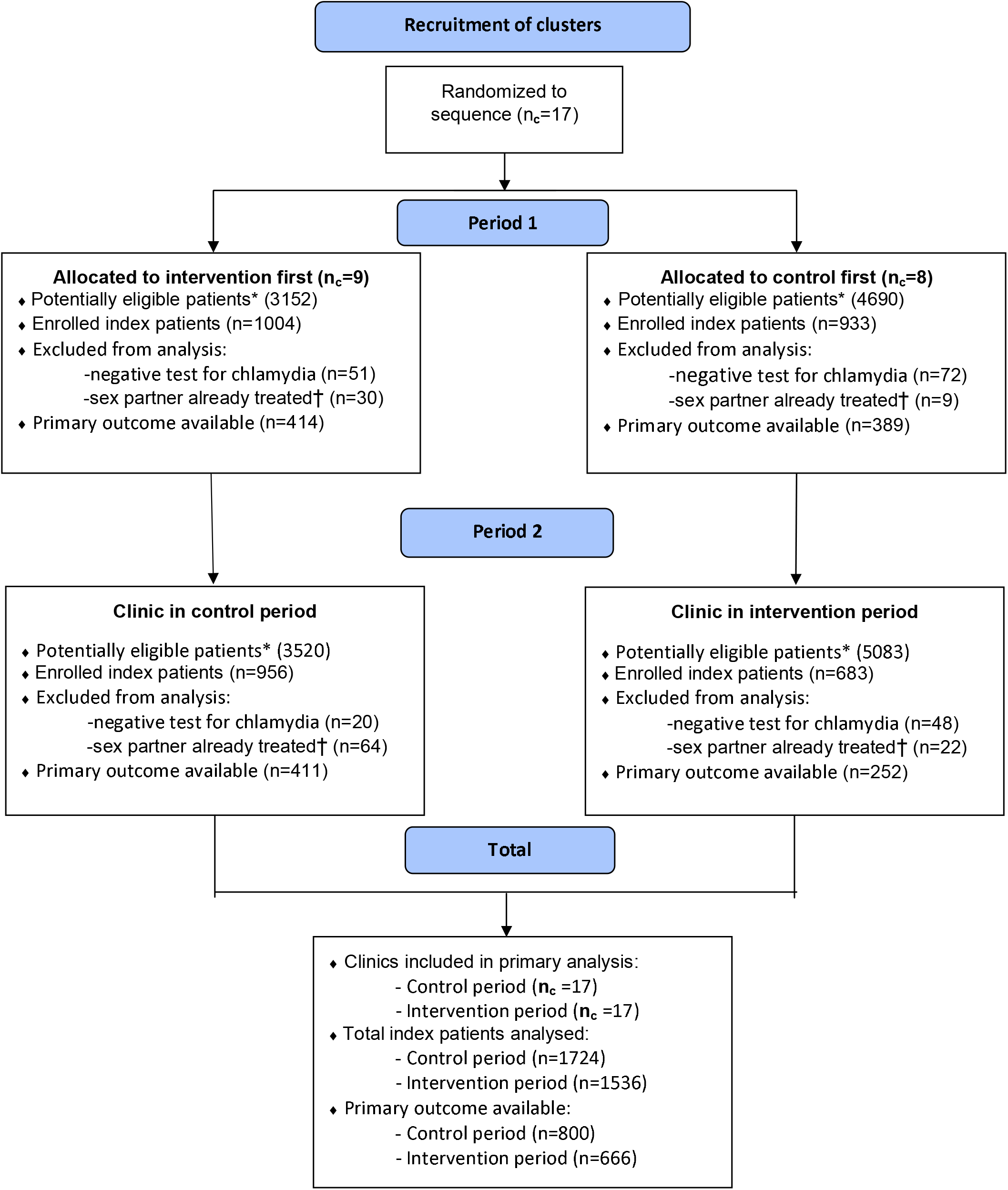
Flow diagram of enrolment by clinic randomisation status and period. *Administrative service data on all chlamydia diagnoses within trial period in non-MSM patients aged ≥16 years not attending as partner notification contact. †All potentially eligible sex partners treated prior to clinic consultation of index patient.

**Figure 2:**
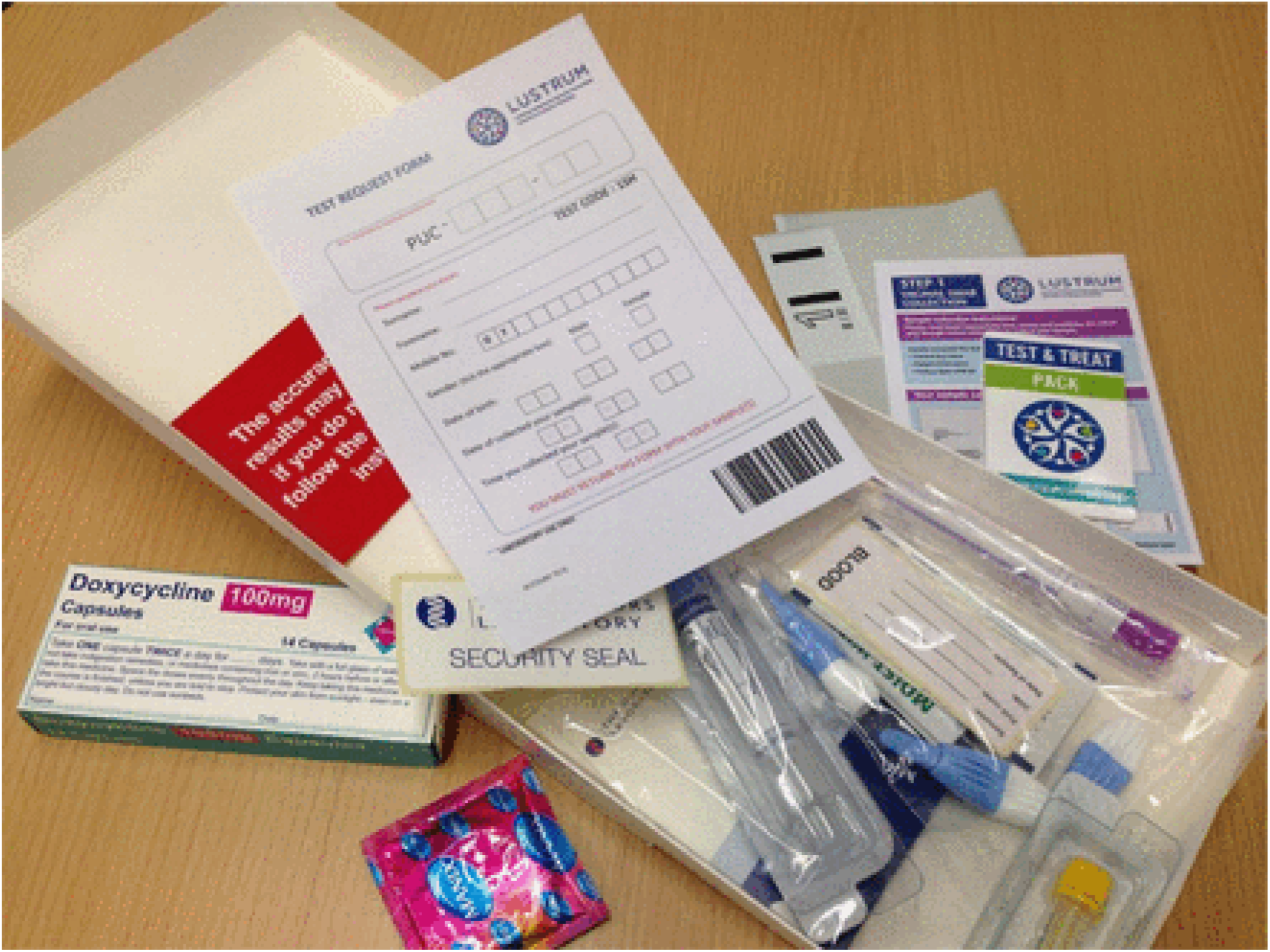
The APT Test and Treat Pack. The APT pack is packaged in a plain white box with no markings or identifiers and contains: antibiotics; condoms; Limiting Undetected Sexually Transmitted infections to RedUce Morbidity TEST & TREAT leaflet; vulvo-vaginal swab kit or urine sampling kit; blood sampling kit; instruction leaflet for sampling kits; test request form for sample processing; prepaid return post envelope; security seal sticker; attention card

**Table 1:** Baseline characteristics of the index patients (IP)

| Variable |  | Control period, *n* (%) or median (IQR) | Intervention period, *n* (%) or median (IQR) |
| --- | --- | --- | --- |
| Number of IPs |  | 1724 | 1536 |
| **Socio-demographic factors** |  |  |  |
| Age | Years (median (IQR, range)) | 24 (21-28, 17-62) | 24 (21-28, 16-72) |
| IP sex at birth* | Male | 547 (32) | 522 (34) |
|  | Female | 1177 (68) | 1014 (66) |
|  | Other | 0 (0) | 0 (0) |
| Enrolment of IP based on | Diagnosed chlamydia | 1678 (97) | 1506 (98) |
|  | PID | 7 (0.4) | 1 (0.06) |
|  | Cervicitis | 0 (0) | 0 (0) |
|  | NGU | 37 (2.1) | 27 (1.7) |
|  | Epididymo-orchitis | 2 (0.12) | 2 (0.13) |
| Ethnicity | White British or Irish | 829 (48) | 707 (46) |
|  | White other | 199 (12) | 181 (12) |
|  | Black /Black British | 368 (21) | 377 (25) |
|  | Asian/British Asian | 100 (6) | 92 (6) |
|  | Mixed ethnicity | 193 (11) | 134 (9) |
|  | Other ethnicity | 35 (2) | 45 (3) |
| **Sex partners per IP** |  |  |  |
| Sex partners last 12 months | Count (median (IQR, range)) | 2 (1-3, 1-100) | 2 (1-4, 1-60) |
| New sex partners last 12 months | Count (median (IQR, range)) | 2 (1-3, 0-99) | 1 (1-3, 0-50) |
| Sex partners in 1/3/6 month look-back† | Count (median (IQR, range)) | 2 (1-2, 1-25) | 1 (1-2, 1-39) |
| Sex partners included in analysis | Count (median (IQR, range)) | 1 (1-2, 1-20) | 1 (1-2, 1-10) |

IQR, interquartile range. \*This was the same as current gender identity in all IPs in the primary analysis. †Dependent on basis for initial enrolment.

**Table 2:**
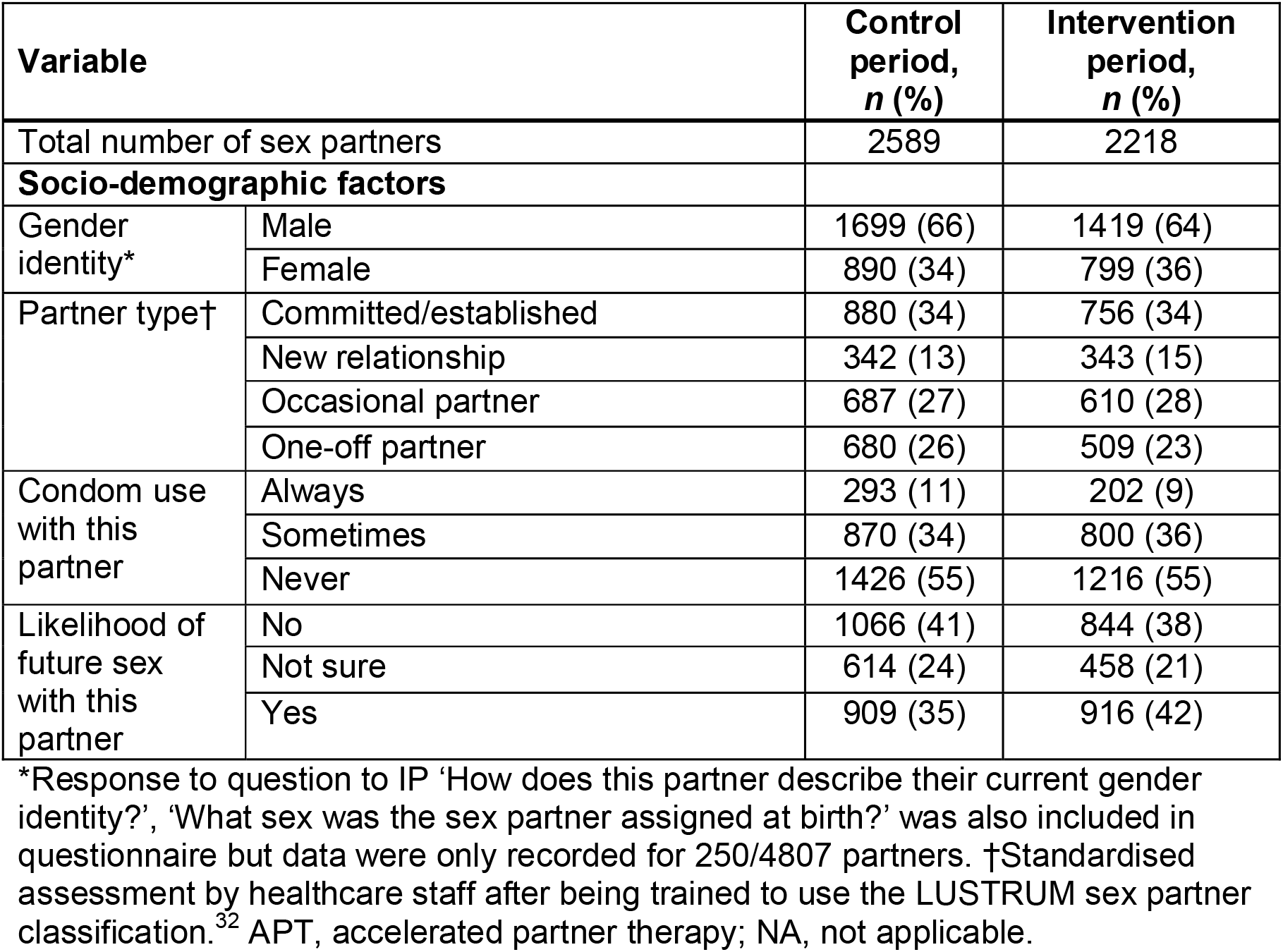
Characteristics of the sex partners (following data provided by index patient)

| Variable |  | Control period, *n* (%) | Intervention period, *n* (%) |
| --- | --- | --- | --- |
| Total number of sex partners |  | 2589 | 2218 |
| **Socio-demographic factors** |  |  |  |
| Gender identity* | Male | 1699 (66) | 1419 (64) |
|  | Female | 890 (34) | 799 (36) |
| Partner type† | Committed/established | 880 (34) | 756 (34) |
|  | New relationship | 342 (13) | 343 (15) |
|  | Occasional partner | 687 (27) | 610 (28) |
|  | One-off partner | 680 (26) | 509 (23) |
| Condom use with this partner | Always | 293 (11) | 202 (9) |
|  | Sometimes | 870 (34) | 800 (36) |
|  | Never | 1426 (55) | 1216 (55) |
| Likelihood of future sex with this partner | No | 1066 (41) | 844 (38) |
|  | Not sure | 614 (24) | 458 (21) |
|  | Yes | 909 (35) | 916 (42) |

\*Response to question to IP 'How does this partner describe their current gender identity?', 'What sex was the sex partner assigned at birth?' was also included in questionnaire but data were only recorded for 250/4807 partners. †Standardised assessment by healthcare staff after being trained to use the LUSTRUM sex partner classification.<sup>32</sup> APT, accelerated partner therapy; NA, not applicable.

In the control and intervention phases, 800 (46%) and 666 (43%) of index patients returned a sample for *C. trachomatis* testing at 12-24 weeks after initial contact tracing consultation. Of those tested 53/800 (6.6%) in the control phase and 31/666 (4.7%) in the intervention phase had a positive result, adjusted OR (aOR) 0.66 (95% CI 0.41–1.04, p=0.07) (Table 3). Analysis after ‘missing at random’ multiple imputation was consistent with the observed data analysis, but varying our assumptions led to stronger effect estimates both if those who did not return a test were thought more likely to be positive than those who did (aOR 0.58, 0.36– 0.92) and for the converse (0.57, 0.37–0.88).

**Table 3:** Effect of ‘offer of APT’ on outcome measures at level of index patient (IP)

| Outcome measures | Control period | Intervention period |  |  |
| --- | --- | --- | --- | --- |
|  | *n (%)#* | *n (%)#* | OR (95% CI); *P*-value | aOR (95% CI); *P*-value |
| Number of IPs | 1724 | 1536 |  |  |
| **Primary outcome** |  |  |  |  |
| IP chlamydia test 12-24 weeks (observed data) |  |  |  |  |
| Positive | 53 (6.6) | 31 (4.7) | 0.67 (0.42–1.06); 0.08 | 0.66 (0.41–1.04); 0.07 |
| Negative | 747 (93.4) | 635 (95.3) | — | — |
| No test* | 924 | 870 | Excluded* | Excluded* |
| IP chlamydia test 12-24 weeks (MAR MI) |  |  |  |  |
| Positive | 116 (6.7) | 73 (4.8) | 0.67 (0.40–1.14); 0.14 | 0.67 (0.39–1.14); 0.14 |
| Negative | 1608 (93.3) | 1463 (95.2) | — | — |
| IP chlamydia test 12-24 weeks (MNAR MI: $\delta = \log_e(0.5)$) | | | | |
| Positive | 154 (8.9) | 86 (5.6) | 0.58 (0.36–0.92); 0.02 | 0.58 (0.36–0.92); 0.02 |
| Negative | 1570 (91.1) | 1450 (94.4) | — | — |
| IP chlamydia test 12-24 weeks (MNAR MI: $\delta = \log_e(2)$) | | | | |
| Positive | 98 (5.7) | 55 (3.6) | 0.57 (0.38–0.88); 0.01 | 0.57 (0.37–0.88); 0.01 |
| Negative | 1626 (94.3) | 1481 (96.4) | — | — |
| **Secondary outcome** |  |  |  |  |
| ≥1 sex partner treated for chlamydia (observed data) |  |  |  |  |
| Yes† | 760 (84.6) | 775 (88.0) | 1.25 (0.94–1.64); 0.12 | 1.27 (0.96–1.68); 0.10 |
| No‡ | 138 (15.4) | 106 (12.0) | — | — |
| Not known* | 826 | 655 | Excluded* | Excluded* |
| ≥1 sex partner treated for chlamydia (MAR MI) |  |  |  |  |
| Yes | 1452 (84.2) | 1344 (87.5) | 1.29 (0.94–1.77); 0.12 | 1.30 (0.94–1.81); 0.12 |
| No¶ | 272 (15.8) | 192 (12.5) | — | — |
| ≥1 sex partner notified (observed data) |  |  |  |  |
| Yes† | 1185 (97.3) | 1123 (97.7) | 1.17 (0.69–1.97); 0.56 | 1.18 (0.70–2.00); 0.54 |
| No‡ | 33 (2.7) | 27 (2.3) | — | — |
| Not known* | 506 | 386 | Excluded* | Excluded* |

\*Considered missing and not included in model estimation. †Determined by follow-up interview with IP, or return of APT self-test kits within 30 days. ‡Includes a mixture of 'no' and 'unknown' treatment outcomes for sex partners listed for a single IP. ¶Mixture of 'no' and 'unknown' treatment outcomes for sex partners listed for a single IP treated as observed 'No' rather than imputed. #Mean average value across imputations reported where relevant. aOR, adjusted odds ratio; MAR, missing at random; MI, multiple imputation; MNAR, missing not at random; OR, odds ratio.

The proportion of index patients with one or more sex partner notified was 1123/1150 (97.7%) in the intervention phase and 1185/1218 (97.3%) in the control phase (aOR 1.18, 95% CI 0.70-2.00, p=0.54), whilst the proportion of all partners notified was 95% in both phases (aOR 0.80, 95% CI 0.49-1.29, p=0.35). The proportion of index patients with one or more sex partners treated 2-4 weeks after the contact tracing consultation was 760/898 (84.6%) in the control phase and 775/881 (88.0%) in the intervention phase, where recorded (aOR 1.27, 95% CI 0.96–1.68, p=0.10) (Table 3).

However, of all sex partners, only 859/2589 (33.2%) were known to be treated by 2-4 weeks in the control phase (400/880 (45.5%) committed/established, 151/342 (44.2%) new, 175/687 (25.5%) occasional and 133/680 (19.6%) one-off) and 842/2218 (38.0%) in the intervention phase (400/756 (52.9%) committed/established, 182/343 (53.1%) new, 162/610 (26.6%) occasional and 98/509 (19.3%) one-off) (Table 4). Overall, less than one fifth (231/1189) of reported one-off partners were known to be treated.

**Table 4:**
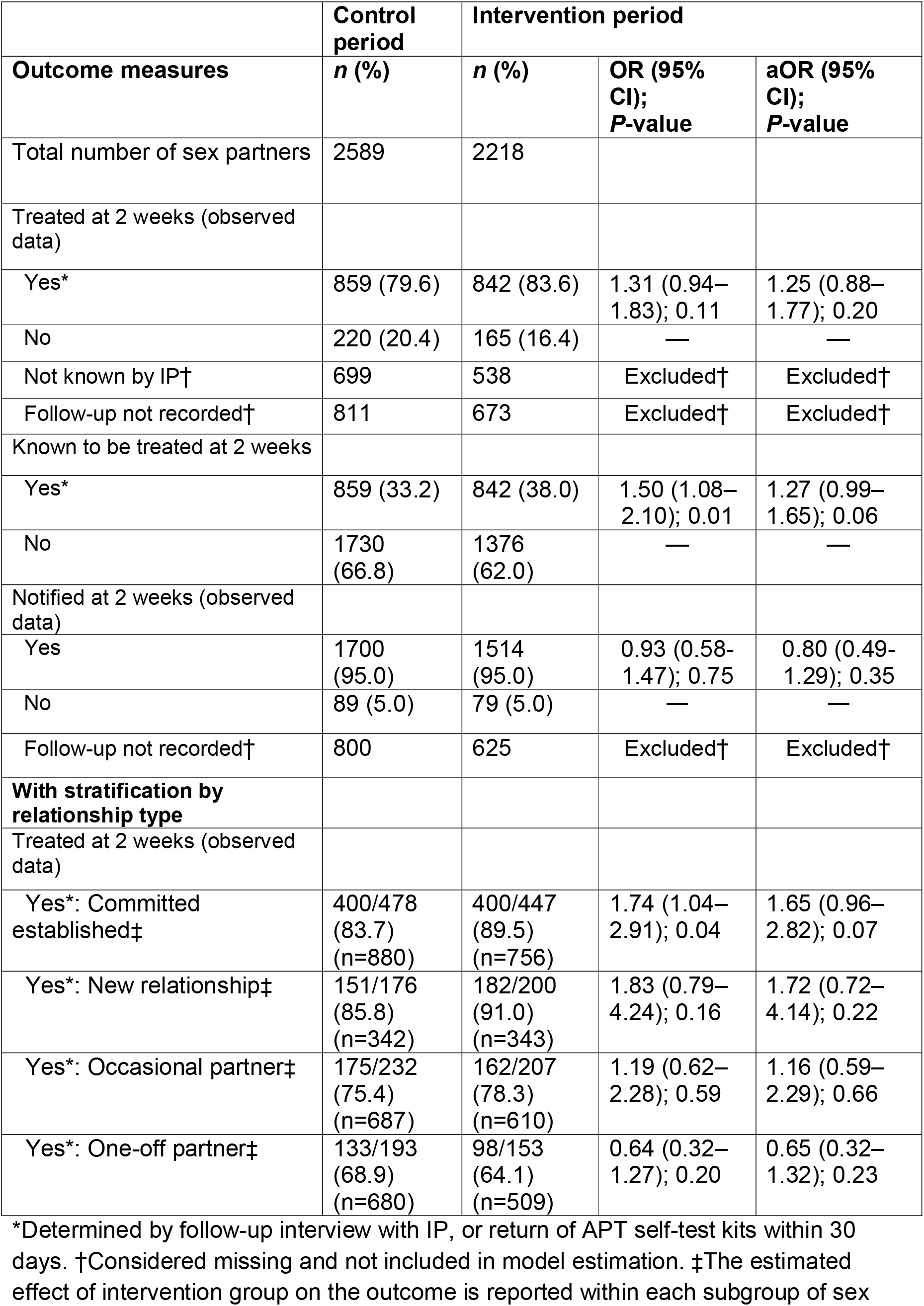
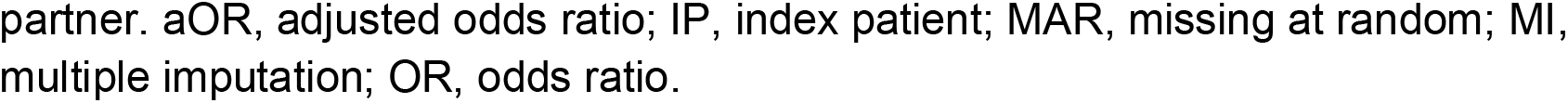
Effect of ‘offer of APT’ on outcome measures at level of sex partner

|  | **Control period** | **Intervention period** |  |  |
| --- | --- | --- | --- | --- |
| **Outcome measures** | **n (%)** | **n (%)** | **OR (95% CI); P-value** | **aOR (95% CI); P-value** |
| Total number of sex partners | 2589 | 2218 |  |  |
| Treated at 2 weeks (observed data) |  |  |  |  |
| Yes* | 859 (79.6) | 842 (83.6) | 1.31 (0.94–1.83); 0.11 | 1.25 (0.88–1.77); 0.20 |
| No | 220 (20.4) | 165 (16.4) | — | — |
| Not known by IP† | 699 | 538 | Excluded† | Excluded† |
| Follow-up not recorded† | 811 | 673 | Excluded† | Excluded† |
| Known to be treated at 2 weeks |  |  |  |  |
| Yes* | 859 (33.2) | 842 (38.0) | 1.50 (1.08–2.10); 0.01 | 1.27 (0.99–1.65); 0.06 |
| No | 1730 (66.8) | 1376 (62.0) | — | — |
| Notified at 2 weeks (observed data) |  |  |  |  |
| Yes | 1700 (95.0) | 1514 (95.0) | 0.93 (0.58–1.47); 0.75 | 0.80 (0.49–1.29); 0.35 |
| No | 89 (5.0) | 79 (5.0) | — | — |
| Follow-up not recorded† | 800 | 625 | Excluded† | Excluded† |
| **With stratification by relationship type** |  |  |  |  |
| Treated at 2 weeks (observed data) |  |  |  |  |
| Yes*: Committed established‡ | 400/478 (83.7) (n=880) | 400/447 (89.5) (n=756) | 1.74 (1.04–2.91); 0.04 | 1.65 (0.96–2.82); 0.07 |
| Yes*: New relationship‡ | 151/176 (85.8) (n=342) | 182/200 (91.0) (n=343) | 1.83 (0.79–4.24); 0.16 | 1.72 (0.72–4.14); 0.22 |
| Yes*: Occasional partner‡ | 175/232 (75.4) (n=687) | 162/207 (78.3) (n=610) | 1.19 (0.62–2.28); 0.59 | 1.16 (0.59–2.29); 0.66 |
| Yes*: One-off partner‡ | 133/193 (68.9) (n=680) | 98/153 (64.1) (n=509) | 0.64 (0.32–1.27); 0.20 | 0.65 (0.32–1.32); 0.23 |

\*Determined by follow-up interview with IP, or return of APT self-test kits within 30 days. †Considered missing and not included in model estimation. ‡The estimated effect of intervention group on the outcome is reported within each subgroup of sex
partner. aOR, adjusted odds ratio; IP, index patient; MAR, missing at random; MI, multiple imputation; OR, odds ratio.

A total of 1536 index patients with 2218 partners were enrolled in APT intervention phases, but APT could not be offered by the clinic in 81/2218 of these. The index patient selected APT for 305/2137 (14.3%) partners when available (Table 5). Of these, 166/305 (54%) were established/committed partners, 85/305 (29%) new relationships, 45 (15%) occasional and 9 (3%) one-off partners. Common reasons for an index patient declining APT included: preference for face-to-face conversation 400/1832 (21.8%), the partner was already in clinic 388/1832 (21.2%), the patient was unwilling to engage with the partner 206/1832 (11.2%), the patient preferred the partner to attend clinic 202/1832 (11.0%), or the partner was overseas 150/1832 (8.2%).

**Table 5:**
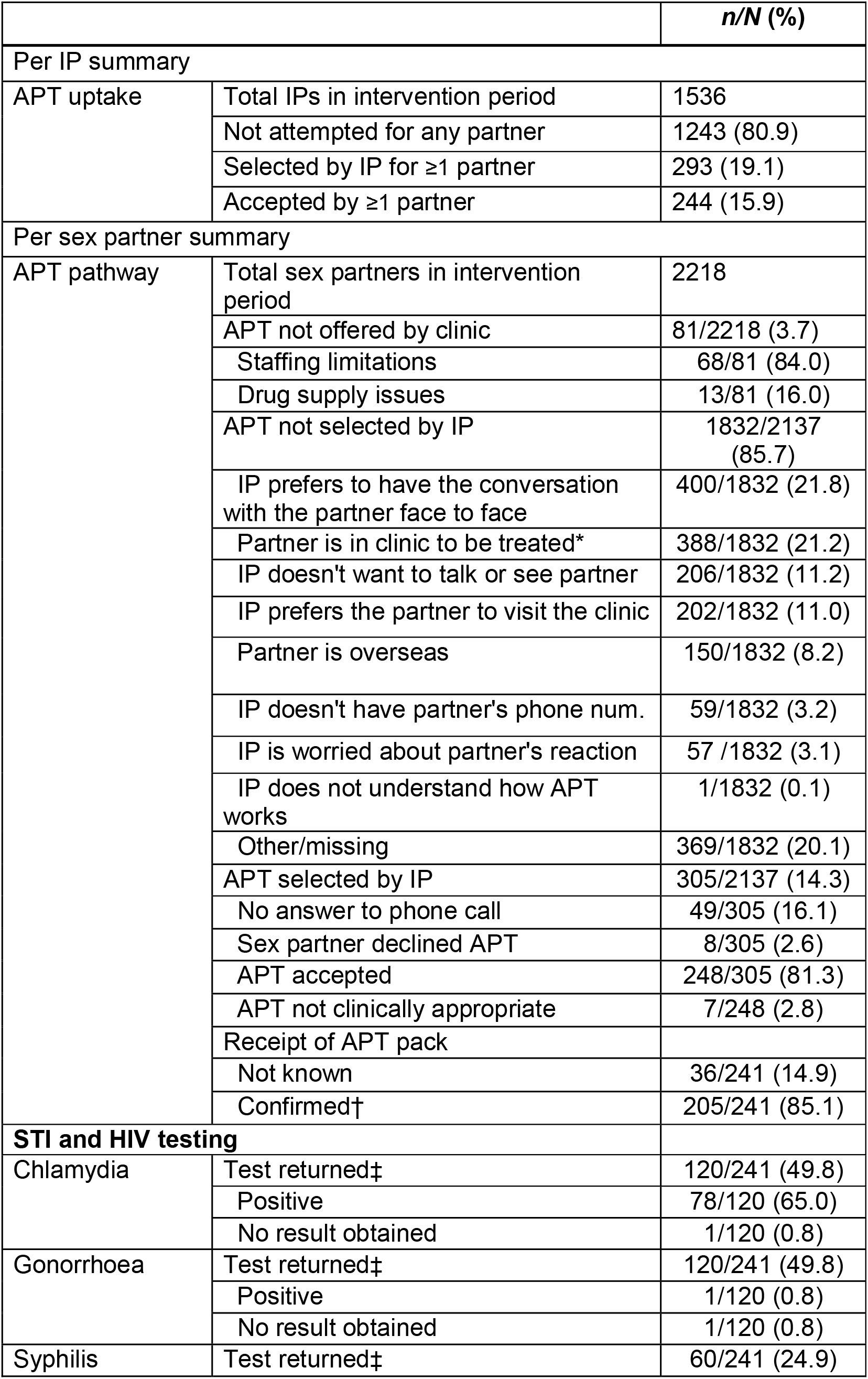
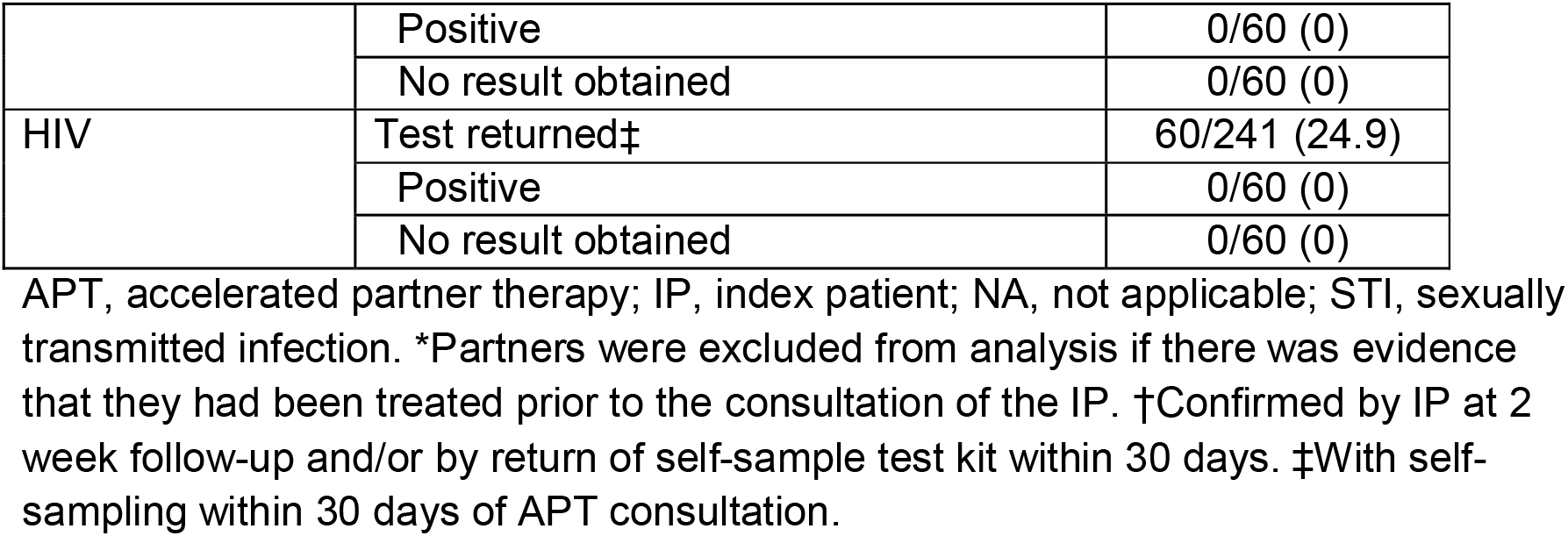
Summary of APT uptake and STI and HIV testing amongst sex partners during intervention phase

**Table 6:** Sensitivity analysis of the effect of ‘offer of APT’ on outcome measures at level of index patient (IP), excluding data from 6/17 clinics with proportion of IPs with APT accepted for at least one partner below 15%.

| Outcome measures | Control period | Intervention period |  |  |
| --- | --- | --- | --- | --- |
|  | *n* (%) | *n* (%) | OR (95% CI); *P*-value | aOR (95% CI); *P*-value |
| Number of IPs | 828 | 586 |  |  |
| **Primary outcome** |  |  |  |  |
| IP chlamydia test 12-24 weeks (observed data) |  |  |  |  |
| Positive | 30 (7.8) | 12 (5.0) | 0.59 (0.29–1.18); 0.12 | 0.56 (0.28–1.13); 0.09 |
| Negative | 357 (92.2) | 229 (95.0) | — | — |
| No test* | 441 | 345 | Excluded* | Excluded* |
| **Secondary outcome** |  |  |  |  |
| ≥1 sex partner treated for chlamydia (observed data) |  |  |  |  |
| Yes† | 342 (83.8) | 318 (90.1) | 1.72 (1.09–2.73); 0.02 | 1.72 (1.08–2.72); 0.02 |
| No‡ | 66 (16.2) | 35 (9.9) | — | — |
| Not known* | 420 | 233 | Excluded* | Excluded* |

\*Considered missing and not included in model estimation. †Determined by follow-up interview with IP, or return of APT self-test kits within 30 days. ‡Includes a mixture of ‘no’ and ‘unknown’ treatment outcomes for sex partners listed for a single IP. aOR, adjusted odds ratio; OR, odds ratio.

Once the index patient had selected APT, care of the sex partner largely followed all specified steps (Table 5), but 49/305 (16%) sex partners could not be reached by telephone, 8/305 (3%) partners declined APT and 7/305 (2%) were transferred into face-to-face clinical care. Of 241 partners sent APT packs, 183 (76%) were male, 120/241 (49.8%) returned chlamydia and gonorrhoea testing samples, of which 78/119 (65.5%) were positive for chlamydia (no result obtained for one returned sample), but only 60/241 (24.9%) returned HIV and syphilis samples (all negative) (Table 5).

We conducted a ‘per protocol’ analysis (not pre-specified) based on index patients who chose APT that was accepted for one or more sex partner. Of 106 such index patients that tested for chlamydia at 12-24 weeks, only 2 (1.9%) were positive, compared to 6.6% (53) in the control arm, aOR 0.26 (95% CI 0.06-1.07) (Table S3). Test positivity was 5.2% (29) in index patients not selecting APT or whose partners refused.

There were seven adverse events reported during the trial, all were assessed as low severity by the trial management group and only one had direct implications for analysis. These were all reported to the Trial Steering Committee members who concluded there were no clinically significant harms to patients.

### Time to treatment

There were insufficient data to compare times to sex partner treatment because, although index patients were aware that a partner had been treated, it was unusual to know exactly when treatment occurred.

### Economic evaluation

Full details of the cost consequence analysis are provided in supplementary appendices. Briefly, the total contact tracing cost for the index patient was £71.26 for the control phase, and £91.23 and £74.83 for the intervention phase with and without APT respectively. The cost differences were mostly driven by the costs associated with the estimated duration of the initial consultation. The results suggest the APT strategy is more costly and also more effective in preventing reinfection of index patients. For the sex partners, the total contact tracing cost was £33.17 in the intervention phase with APT compared to £39.58 in the control phase, if we assume that sex partners only returned samples for chlamydia testing. However, if we assume that some sex partners had an additional test for HIV or syphilis, the costs increased to £40.12 (for the APT users) versus £46.53 (for standard contact tracing). This analysis presents preliminary results for the intermediate outcome of reinfections avoided. A full economic evaluation based on a population-based chlamydia transmission model comparing the cost-effectiveness of APT with standard PN in terms of major outcomes averted and quality-adjusted life years gained is reported elsewhere.^29^

### Process Evaluation

Detailed process evaluation findings are reported elsewhere.^28^ Briefly, clinics operationalised the trial in different ways; some aimed to offer APT to all potentially eligible patients while others only managed to offer APT to patients attending on certain days of the week when staff with trial responsibility were present. Staff found that RELAY enhanced their ability to document contact tracing processes and outcomes because it was user-friendly and intuitive to use. Index patients commonly reported that APT was only suitable for certain types of sex partner, particularly committed/established relationships and was not appropriate for relationships with a lower emotional connection (e.g. one-off partners), which largely supports trial findings (Table S2). However, healthcare professionals did not always offer APT owing to multiple and diverse clinic pressures, including a lack of time to create the required RELAY entry as this was additional to documenting outcomes in their clinical notes. Also, some sex partners accompanied the index patients when they attended for treatment or had already accessed face-to-face care. Nevertheless, those who chose APT felt it worked well and helped partners overcome barriers to attending in person. Most sex partners received APT packs directly from the index patient within a day of consultation. Some sex partners took the treatment immediately but waited until treatment was completed to use the self-sampling kits as a “test of cure”. Some sex partners reported difficulties in blood sampling (finger-prick) and some did not understand the rationale for routine “full check-up” testing for STIs other than chlamydia, although this was explained during their consultation. In addition, hospital electronic data communication constraints meant that most clinics were unable to provide the sex partners a direct link to view the short videos we had created to assist sex partner engagement with APT and the packs in particular. These factors could explain lower return of blood samples for HIV and syphilis testing than chlamydia and gonorrhoea samples, which are done as a combined test on urine/vulvo-vaginal swabs.

## Discussion

### Principal findings

The offer of APT in the intervention group resulted in a likely reduction in the proportion of people with repeat chlamydia infection (4.7% vs. 6.6%, aOR 0.66, 95% CI 0.41–1.04, p=0.07) and a likely increase in the proportion of index cases with at least one partner treated (88.0% vs. 84.6%, aOR 1.27, 95% CI 0.96–1.68, p=0.10). Overall, 293/1536 (19.1%) of index patients chose APT for a total of 305 partners, of whom 248 accepted. The APT intervention cost slightly more than standard contact tracing for the index patient, but partner treatment and testing were cheaper.

### Strengths and weaknesses

Strengths of the trial were that we developed the APT intervention following a detailed stepwise framework for complex interventions^20,21,41,42^ and measured the primary outcome with a biological marker of chlamydia infection, as recommended for evaluations of contact tracing strategies.^12^ The APT strategy was theory-informed^43^ and delivered with good/high fidelity^28^ by trained healthcare professionals in a variety of sexual health clinic settings. APT was delivered as a clinic-wide, low-risk intervention, with ethical approval at the service level, using a novel GDPR-compliant process.^40,44^ Recognising the contribution of different types of partnership to STI transmission,^45^ we also examined the effects of APT using a novel classification of sex partner type, which we developed as part of the LUSTRUM programme.^32^

A weakness of the trial was that the frequency of chlamydia infection at the time of repeat testing (6.6% at 12-16 weeks in the control arm) was lower than in the studies on which we based our sample size calculation (10-25%),^11,27,35,46^ reducing the power of the trial. There are several possible explanations for this; 1) improvements in health outcomes over time may have reduced repeat positivity in the population under study, 2) by excluding index patients who only reported uncontactable partners (for whom contact tracing is impossible), we excluded those most likely to have a repeat positive chlamydia test, 3) the cross-over design might have resulted in lower community transmission in both arms and thereby reduced incidence/reinfection in both. APT uptake itself was not a part of the power calculations. Although APT was not always offered, we expected more index patients to choose it when it was made available.^20,21^ Our primary outcome was based on the results of repeat chlamydia tests, which were taken up by slightly less than half the index patients. However, we investigated the potential for bias through multiple imputation under different assumptions, which showed consistent results.

The process evaluation suggested that some of the trial procedures reduced the offering of APT as an option. Despite the pragmatic trial design, which was intended to evaluate APT under real-life clinical conditions, the trial generated additional administrative work, with the need to create a record on RELAY even if the patient did not accept the APT intervention.^28^ Since the software recorded the data and outcomes needed to evaluate APT and usual care, the efficacy of the intervention could not be established without the RELAY system. As a result, many clinics designated only certain sessions each week to recruit to the trial and we saw similar enrolment rates in intervention and control phases and little difference in the characteristics of index patients enrolled between the two phases. Consequently, only a small proportion (roughly one in five) of potentially eligible index patients were enrolled and we accept that selection bias may have affected the trial.

### Strengths and weaknesses in relation to other studies

The evidence from this trial, about the effectiveness of APT for a biological outcome and the inclusion of comprehensive STI and HIV testing, adds to the preliminary evidence from previous studies.^20,21^ Uptake of APT in earlier studies appeared to be associated with how the intervention was operationalised in individual clinics^20^ and varied between 40-80%. In this trial, index patient uptake of APT also differed between clusters (Table S1), largely influenced by the level of enthusiasm for the trial in different settings but also possibly due to changes in how clinics provided usual care over time, such as by encouraging index patients to bring their sex partners with them when they attended for treatment (Table S2).

APT is an adaptation, for the UK setting, of EPT.^47^ In a systematic review, EPT resulted in lower proportions of index cases with repeated curable STIs (any of gonorrhoea, chlamydia or trichomoniasis) than simple patient referral contact tracing.^12^ Golden et al.^24^ used a randomised step-wedge design to evaluate EPT in Washington State, United States of America, and found some evidence of lower chlamydia positivity and gonorrhoea incidence at the population level. There are important differences between the UK and American settings and between APT and EPT. Firstly, baseline and repeat infection rates were considerably higher in US studies than in our trial and pre-existing contact tracing outcomes were poorer than those routinely achieved in the UK. Secondly, EPT trials did not include sex partner STI and HIV testing, so sex partners did not receive contact tracing services. In our trial, almost two thirds of sex partners who returned a sample had a positive chlamydia test result. Onward contact tracing outside the trial might have wider, but unmeasured, effects on community transmission. Thirdly, APT limits treatment to chlamydia because there is no recommended first line oral treatment for *N. gonorrhoeae*. Although EPT guidance still allows oral cefixime for gonorrhoea, an update in 2021 suggests that providers should limit treatment to people who cannot access prompt clinical evaluation and treatment.^23^

### Meaning of the study

APT is an acceptable and cost-saving intervention which could be added to the menu of contact tracing options, allowing sex partners to receive treatment safely after exposure to chlamydia without the need for a clinic appointment. In our trial, APT appeared to reduce the proportion of index cases with repeat chlamydia infection and increased the proportion with at least one sex partner treated, when compared with standard PN. We attribute the modest effect sizes, in part, to the smaller than expected numbers of index patients choosing it for their partners. It is also possible that the use of RELAY, which appealed to clinic staff, systematically enhanced contact tracing processes and outcome recording in usual care, reducing any difference associated with APT itself.

When implemented into routine services, the trial-related administrative work, which was a barrier to enrolment (although agreed by the clinics prior to involvement in the trial) and to the offer of APT, would not exist. This in itself may increase both offer and uptake of APT because staff might offer it more assertively if there is no perceived additional administrative burden. Different types of sexual partner contribute differentially to onward transmission of STIs.^48^ In almost all instances where index cases chose APT, they did so for an established or ongoing partner, for whom evidence shows that routine PN approaches are most successful. However, in the parallel mathematical modelling studies, the base-case results, showed that APT is both less costly and more effective in terms of major outcomes averted (MOA) and quality adjusted life years (QALYs) than standard contact tracing, implying that APT is cost-saving.^29^ In contrast, one-off partnerships made up only one in thirty APT decisions, although these accounted for one in five partnerships. This is noteworthy as one-off partnerships are likely to contribute disproportionately to transmission within the population.^48^

Almost half of sex partners accepting APT returned a sample (urine or vulvo-vaginal swab) for chlamydia and gonorrhoea testing, which was a much higher proportion than in our earlier feasibility study.^20^ Only one in four, however, returned samples for HIV and syphilis testing. In contrast, almost all sex partners who attend sexual health services in person receive testing for venepuncture for serum HIV and other blood borne viruses depending on risk, and syphilis serology.

### Implications for clinical practice, service design and future research

In the UK, the coronavirus disease 2019 (Covid-19) pandemic has accelerated the shift to remote, self-managed healthcare. APT is a cost-saving approach^29^ that contains all recommended elements of sex partner management,^17^ and uptake might increase in a post-Covid-19 environment because of increased familiarity with self-sampling, self-testing and contact tracing as well as the rationale for making individual health decisions for both personal and public benefit. Sexual health services should therefore start to integrate APT into their usual contact tracing practices, where it should be promoted for index patients with established / ongoing sex partners, accompanied by research focussing on normalisation, scale up and skills acquisition. Comprehensive clinician training resources are available from the authors.

However, the well-described, long-term pressures on UK sexual health services^49^ will make it hard for services to provide flexible models of care that allow immediate, and possibly unscheduled, assessment of the sex partner. APT will need to be audited alongside all other contact tracing approaches, so data collection practices, including the recording of partnership type, should be established now, ahead of the next national audit of STI contact tracing which will pilot the new classification of partnership types, which we used in the trial.^32^ More work is needed to increase the uptake of self-sampling for STIs as part of APT, so that opportunities for screening and control of syphilis, HIV and other blood borne viruses amongst those at higher risk of infection are not lost. In addition, the potential harms of APT should continue to be assessed, since “blanket” epidemiologic treatment for sex partners of people with chlamydia, in the absence of a positive test result (also common practice in standard contact tracing) leads to over-use of antibiotics.^26^

More broadly, we need to consider the partners who are less likely to be reached by APT, such as one-off partners with whom future sex is not anticipated. Although not a risk to the index patient, they are likely to make an important contribution to community transmission. Further research is needed to improve PN options for other population-level risk groups, including men who have sex with men, trans and gender-diverse people at higher risk of STIs, HIV and other blood borne viruses.

In summary, APT can be safely offered as a cost-saving contact tracing option for heterosexual people with *C. trachomatis* and might reduce the risk of repeat infection. To maximise the impact of APT, there needs to be a focus on increasing uptake. This will need health care professionals to promote APT for emotionally connected sex partners where future sex is likely, and flexibility in clinic capacity and work flow to accommodate immediate sex partner management during the index patient’s attendance. The latter will only be achieved by a renewed emphasis on prevention of transmission at national level.

## Supporting information

Supplemental Table 1: CONSORT checklist

## Data Availability

The trial dataset is available from UCL data repository

https://rdr.ucl.ac.uk/articles/dataset/LUSTRUM_Accelerated_partner_therapy_APT_cross-over_cluster_randomised_controlled_trial_data/14724669

## Data availability

The trial dataset is available:

## Acknowledgements

The authors would like to thank the following: (1) Trial Steering Committee: Simon Barton (chair), Robbie Currie, David Crundwell, Artemis Koukounari, Lynis Lewis, Alec Miners, Emmanuel Rollings-Kamara, Rachel Shaw, Emma Thompson. (2) Data Monitoring Committee: Simon Barton, David Crundwell, Rebecca Turner, Artemis Koukounari. (3) All members of the LUSTRUM Patient and Public Involvement Group. (4) Participating centres: Barking, Havering and Redbridge University Hospitals NHS Trust (A. Umaipalan), Barts Health NHS Trust (V. Apea), Buckinghamshire Healthcare NHS Trust (J. Sherrard), Chelsea and Westminster Hospital NHS Foundation Trust (C. Evans), Croydon Health Services NHS Trust (D. Phillips), Manchester University NHS Foundation Trust (G. Schembri), Midlands Partnership NHS Foundation Trust (J. Dhar), NHS Greater Glasgow and Clyde (R. Nandwani), Northamptonshire Healthcare NHS Foundation Trust (S. Herbert), Royal Berkshire NHS Foundation Trust (F. Chen), Royal Bournemouth and Christchurch Hospitals NHS Foundation Trust (K. Schroeder), Solent NHS Trust (R. Patel), University Hospitals Birmingham NHS Foundation Trust (J. Ross). The authors would also like to thank epiGenesys who developed the RELAY software; Soazig Clifton (support with ethics submission); Jo Gibbs (support on RELAY development); and all of the staff, management and patients involved in the study at all the participating sexual health clinics.

## Additional descriptive data summaries

Cross-tabulation of ‘likelihood of future sex’ and ‘condom use with this partner’ (as reported by the index patient) for all sex partners included in the analysis:

|  | Condom use |  |  | Total |
| --- | --- | --- | --- | --- |
|  | Always | Sometimes | Never |  |
| Likelihood of future sex |  |  |  |  |
| No | 241 (12.6) | 547 (28.6) | 1122 (58.7) | 1910 |
| Not sure | 127 (11.9) | 361 (33.7) | 584 (54.5) | 1072 |
| Yes | 127 (7.0) | 762 (51.3) | 936 (41.8) | 1825 |
| Total | 495 | 1670 | 2642 | 4807 |

Row-wise percentages are reported.

STI self-test return summary by gender* of sex partner:

Row-wise percentages are reported. STI self-test return summary by gender\* of sex partner:
|  |  | Total | Sex partner:<br>male | Sex partner:<br>female |
| --- | --- | --- | --- | --- |
| APT accepted and sex partner eligible, *n* |  | 241 | 183 | 58 |
| **STI and HIV testing** |  |  |  |  |
| Chlamydia | Test returned† | 120/241 (49.8) | 89/183 (48.6) | 31/58 (53.4) |
|  | Positive | 78/120 (65.0) | 58/89 (65.2) | 20/31 (64.5) |
|  | No result obtained | 1/120 (0.8) | 0/89 (0) | 1/31 (3.2) |
| Gonorrhoea | Test returned† | 120/241 (49.8) | 89/183 (48.6) | 31/58 (53.4) |
|  | Positive | 1/120 (0.8) | 1/89 (1.1) | 0/31 (0) |
|  | No result obtained | 1/120 (0.8) | 0/89 (0) | 1/31 (3.2) |
| Syphilis | Test returned† | 60/241 (24.9) | 38/183 (20.8) | 22/58 (37.9) |
|  | Positive | 0/60 (0) | 0/38 (0) | 0/22 (0) |
|  | No result obtained | 0/60 (0) | 0/38 (0) | 0/22 (0) |
| HIV | Test returned† | 60/241 (24.9) | 38/183 (20.8) | 22/58 (37.9) |
|  | Positive | 0/60 (0) | 0/38 (0) | 0/22 (0) |
|  | No result obtained | 0/60 (0) | 0/38 (0) | 0/22 (0) |

\*Response to question to IP 'How does this partner describe their current gender identity?', 'What sex was the sex partner assigned at birth?' was also included in questionnaire but data were only recorded for 250/4807 partners. †With self-sampling within 30 days of APT consultation.

Time to retest sample for primary outcome of chlamydia retest in index patient: mean 104 days, median 98 days, IQR 90-113 days.

Cross-tabulation of primary outcome of index patient retest result according to the acceptance of APT by at least one sex partner during the intervention period:

|  | Control phase | Intervention phase |  | Total |
| --- | --- | --- | --- | --- |
|  |  | No partners accepting APT | ≥1 partners accepting APT |  |
| Index retest result |  |  |  |  |
| Positive | 53 (3.1) | 29 (2.2) | 2 (0.8) | 84 |
| Negative | 747 (43.3) | 531 (41.1) | 104 (42.6) | 1382 |
| No test | 924 (53.6) | 732 (56.7) | 138 (56.6) | 1794 |
| Total | 1724 | 1292 | 244 | 3260 |

Column-wise percentages are reported.

**Table S1:** Cluster-period level summary of baseline data for index patients and APT uptake

| **Variable** |  | **Control period,**<br>**(median (IQR, range))** | **Intervention period,**<br>**(median (IQR, range))** |
| --- | --- | --- | --- |
| Number of cluster-periods |  | 17 | 17 |
| N IPs per cluster-period |  | 107 (73-139, 19-194) | 84 (34-134, 10-246) |
| **Socio-demographics per cluster-period** |  |  |  |
| IP Age | Mean years | 25.3 (24.3-26.2, 22.6-28.1) | 25.1 (24.0-26.8, 22.7-27.6) |
| IP gender identity | Prop. male | 0.32 (0.29-0.38, 0.19-0.58) | 0.34 (0.28-0.36, 0.18-0.80) |
| Ethnicity | Prop. white British or Irish | 0.56 (0.26-0.80, 0.14-0.89) | 0.59 (0.30-0.80, 0.10-0.83) |
|  | Prop. white other | 0.07 (0.06-0.10, 0-0.31) | 0.08 (0.05-0.12, 0-0.26) |
|  | Prop. Black /Black British | 0.14 (0.04-0.30, 0-0.58) | 0.15 (0.07-0.24, 0.03-0.52) |
|  | Prop. Asian/British Asian | 0.04 (0.02-0.08, 0-0.15) | 0.03 (0.01-0.07, 0-0.15) |
|  | Prop. mixed ethnicity | 0.11 (0.05-0.14, 0.01-0.18) | 0.09 (0.04-0.12, 0.03-0.17) |
|  | Prop. other ethnicity | 0.01 (0-0.02, 0-0.08) | 0.01 (0-0.03, 0-0.10) |
| **Post-enrolment sex partner data** |  |  |  |
| Mean sex partners included in analysis per IP per cluster-period |  | 1.5 (1.4-1.7, 1.0-2.6) | 1.4 (1.2-1.6, 1.0-1.7) |
| Prop. of sex partners with APT selected |  | — | 0.19 (0.10-0.25, 0.03-0.71) |
| Prop. of sex partners accepting APT |  | — | 0.14 (0.08-0.25, 0.02-0.64) |
| Prop. of sex partners known to be treated |  | 0.31 (0.29-0.37, 0.16-0.52) | 0.40 (0.32-0.47, 0.26-0.75) |
| **Outcome data** |  |  |  |
| Prop. of IPs with primary outcome available |  | 0.46 (0.43-0.52, 0.30-0.56) | 0.41 (0.36-0.46, 0.23-0.53) |
| Prop. of IPs with positive retest result* |  | 0.07 (0.04-0.08, 0-0.16) | 0.03 (0-0.06, 0-0.2) |

Median, IQR and range values are given across the cluster-period level values. \*Of those with retest result available. ATP, accelerated partner therapy; IP, index patient.

**Table S2:** Summary of APT uptake and STI and HIV testing amongst sex partners during the intervention phase, stratified by partner type

|  | Total | Committed/<br>established | New<br>relationship | Occasional<br>partner | One-off partner |
| --- | --- | --- | --- | --- | --- |
| **Per sex partner summary of APT pathway** |  |  |  |  |  |
| Total sex partners in intervention period | 2218 | 756 | 343 | 610 | 509 |
| APT not offered by clinic | 81/2218 (3.7) | 26/756 (3.4) | 9/343 (2.6) | 31/610 (5.1) | 15/509 (2.9) |
| Staffing limitations | 68/81 (84.0) | 22/26 (84.6) | 8/9 (88.9) | 24/31 (77.4) | 14/15 (93.3) |
| Drug supply issues | 13/81 (16.0) | 4/26 (15.4) | 1/9 (11.1) | 7/31 (22.6) | 1/15 (6.7) |
| APT not selected by IP | 1832/2137 (85.7) | 564/730 (77.3) | 249/334 (74.6) | 534/579 (92.2) | 485/494 (93.7) |
| IP doesn't have partner's phone number | 59/1832 (3.2) | 6/564 (1.1) | 2/249 (0.8) | 11/534 (2.1) | 40/485 (8.3) |
| Partner is overseas | 150/1832 (8.2) | 55/564 (9.8) | 15/249 (6.0) | 36/534 (6.7) | 44/485 (9.1) |
| IP is worried about partner's reaction | 57 /1832 (3.1) | 21/564 (3.7) | 8/249 (3.2) | 15/534 (2.8) | 13/485 (2.7) |
| IP doesn't want to talk or see partner | 206/1832 (11.2) | 49/564 (8.7) | 15/249 (3.2) | 64/534 (12.0) | 78/485 (16.1) |
| IP prefers to have the conversation with the partner face to face | 400/1832 (21.8) | 97/564 (17.2) | 56/249 (22.5) | 137/534 (25.7) | 110/485 (22.7) |
| IP prefers the partner to visit the clinic | 202/1832 (11.0) | 67/564 (11.9) | 36/249 (14.5) | 54/534 (10.1) | 45/485 (9.3) |
| IP does not understand how APT works | 1/1832 (0.1) | 1/564 (0.2) | 0/249 (0) | 0/534 (0) | 0/485 (0) |
| Partner is in clinic to be treated* | 388/1832 (21.2) | 166/564 (29.4) | 75/249 (30.1) | 95/534 (17.8) | 52/485 (10.7) |
| Other/missing | 369/1832 (20.1) | 102/564 (18.1) | 42/249 (16.9) | 122/534 (22.9) | 103/485 (21.2) |
| APT selected by IP | 305/2137 (14.3) | 166/730 (22.7) | 85/334 (25.4) | 45/579 (7.8) | 9/494 (1.8) |
| No answer to phone call | 49/305 (16.1) | 20/166 (12.0) | 9/85 (10.6) | 16/45 (35.6) | 4/9 (44.4) |
| Sex partner declined APT | 8/305 (2.6) | 4/166 (2.4) | 1/85 (1.2) | 2/45 (4.4) | 1/9 (11.1) |
| APT accepted | 248/305 (81.3) | 142/166 (85.5) | 75/85 (88.2) | 27/45 (60.0) | 4/9 (44.4) |
| APT not clinically appropriate | 7/248 (2.8) | 3/142 (2.1) | 2/75 (2.7) | 2/27 (7.4) | 0/4 (0) |
| Receipt of APT pack |  |  |  |  |  |
| Not known | 36/241 (14.9) | 20/139 (14.4) | 9/73 (12.3) | 6/25 (24.0) | 1/4 (25.0) |
| Confirmed† | 205/241 (85.1) | 119/139 (85.6) | 64/73 (87.7) | 19/25 (76.0) | 3/4 (75.0) |

| **STI and HIV testing** |  |  |  |  |  |  |
| --- | --- | --- | --- | --- | --- | --- |
| Chlamydia | Test returned | 119/241 (49.4) | 65/139 (46.8) | 42/73 (57.5) | 10/25 (40.0) | 2/4 (50.0) |
|  | Positive | 78/119 (65.5) | 45/65 (69.2) | 24/42 (57.1) | 9/10 (90.0) | 0/2 (0) |
| Gonorrhoea | Test returned | 119/241 (49.4) | 65/139 (46.8) | 42/73 (57.5) | 10/25 (40.0) | 2/4 (50.0) |
|  | Positive | 1/119 (0.8) | 1/65 (1.5) | 0/42 (0) | 0/10 (0) | 0/2 (0) |
| Syphilis | Test returned | 60/241 (24.9) | 35/139 (25.2) | 18/73 (24.7) | 6/25 (24.0) | 1/4 (25.0) |
|  | Positive | 0/60 (0) | 0/35 (0) | 0/18 (0) | 0/6 (0) | 0/1 (0) |
| HIV | Test returned | 60/241 (24.9) | 35/139 (25.2) | 18/73 (24.7) | 6/25 (24.0) | 1/4 (25.0) |
|  | Positive | 0/60 (0) | 0/35 (0) | 0/18 (0) | 0/6 (0) | 0/1 (0) |

Data shown as *n/N* (%).
APT, accelerated partner therapy; IP, index patient; NA, not applicable; STI, sexually transmitted infection. \*Partners were excluded from analysis if there was evidence that they had been treated prior to the consultation of the IP. †Confirmed by IP at 2 week follow-up and/or by return of self-sample test kit within 30 days.

**Table S3:** ‘As treated’ effect of ‘APT selected and accepted for at least one sex partner’ on primary outcome of index patient (IP) chlamydia retest at 12-24 weeks

| Outcome measures | Negative | Positive |  |  | No test |
| --- | --- | --- | --- | --- | --- |
|  | *n* (%) | *n* (%) | OR (95% CI); *P*-value | aOR (95% CI); *P*-value | *n* |
| Control period | 747 (93.4) | 53 (6.6) | REF | REF | 924* |
| Intervention period:<br>APT accepted by one<br>or more partner | 104 (98.1) | 2 (1.9) | 0.26 (0.06–1.09); 0.07 | 0.26 (0.06–1.07); 0.06 | 138* |
| Intervention period:<br>APT accepted by no<br>partners or selected<br>for no partners | 531 (94.8) | 29 (5.2) | 0.75 (0.47–1.20); 0.23 | 0.74 (0.46–1.18); 0.21 | 732* |

\*Considered missing and not included in model estimation. aOR, adjusted odds ratio; OR, odds ratio.

## APPENDIX

### Further Statistical Analysis details for Appendix

#### Adjustment for design and baseline factors

We accounted for the structure of the data with random effects for each clinic and for each control/intervention period within each clinic. All analyses included a fixed effect for time period and were also adjusted by the baseline demographic variables for the index patient of age (on continuous scale using 5 knot restricted cubic spline), gender identity at enrolment (female, male; using sex assigned at birth if ‘other’), NGU without definitive chlamydia diagnosis at time of enrolment (these constitute the only symptom-subgroup with a substantial number of patients) and ethnicity (White British or Irish, White other, Black /Black British, Asian/British Asian, Mixed ethnicity, Other ethnicity). These adjustments were pre-specified in the statistical analysis plan (SAP).

Adjusted effect measures were considered the primary effect measures for outcomes analysed according to clinic randomisation status, though unadjusted effect estimates were also reported for completeness.

For outcomes evaluated at the level of ‘sex partner’, analyses were also adjusted for type of sex partner (committed/established, new relationship, occasional partner, one-off partner), condom use (always, sometimes, never) and likelihood of future sex (no, not sure, yes) as reported by the index patient and the total number of sex partners listed by the index patient (1, 2, 3, ≥4).

#### Mixed effects model structure

Both the primary and first secondary (proportion of index patients with one or more partner treated) outcomes are binary and defined at the level of the index patient. The Protocol for the LUSTRUM trial specified that these would be analysed using mixed effects logistic regression models with fixed effects for clinic and intervention condition, together with a random effect to acknowledge the clustering of index patients for each combination of clinic and period (i.e. independent random effects are defined for each of the two periods for each clinic) (model M2).^37^ The model choice specified in the Protocol was based on recommendations for the analysis of trials with continuous outcomes,^37^ however, since then we became aware of a paper evaluating model choices for cluster-randomised crossover trials with binary outcomes;^50^ this study found that a fixed-random model structure (i.e. M2) was susceptible to convergence problems.

Morgan et al. recommend a cluster-level approach to analysis,^50^ but this does not allow for adjustment of individual patient characteristics. The authors found a hierarchical model with random effects for both cluster and cluster-period (and with fixed effect for treatment effect) (H2) to perform well in some scenarios. We carried out simulations, with 1000 datasets generated based on the numbers specified in the LUSTRUM sample size calculations, and confirmed that model H2 would provide appropriate type I error rate and power for the LUSTRUM trial. We therefore used a hierarchical random-random model for the analysis (model H2). This was pre-specified in the SAP.

Our simulations indicated that LRTs showed close to nominal type I error rates for model H2 under the null hypothesis (5.1% and 5.5% for 10% or 25% prevalence of reinfection), whereas the type I error rate was slightly raised when using Wald tests (6.1%, 7.4%). It is known that LRTs are more robust than Wald tests, as the latter rely on stronger assumptions regarding the curvature of the log likelihood function.^51^ We therefore used LRTs for hypothesis testing and the generation of P-values for treatment effects. The intervention effect will be expressed as an odds ratio, with 95% confidence interval calculated using the Wald method.

#### Multiple imputation by chained equations (MICE)

We conducted sensitivity analyses for the primary outcome and sex partner treatment success based on multiple imputation. This first followed a missing at random (MAR) assumption. The term MAR means that outcomes are dependent on the values of the observed data, but not dependent on the values of the missing data. This was conducted using multiple imputation by chained equations (MICE).^52^

The MICE procedure included baseline variables for index patients. It was planned in the SAP that the imputation would also include sex partner level treatment outcomes and characteristics within a single procedure. However, it was not possible to carry out the planned simultaneous multilevel imputation of both missing sex partner-level treatment outcomes and index patient retest outcomes. This was due both to computational issues for the full specified model (low variation in observed sex partner treatment outcomes within each index patient, where multiple partners listed), and lack of convergence to a stable distribution of imputed values even for a simplified version of the model including both partner and index patient-level data. As such, multiple imputation has been conducted only using variables at the level of the index patient, with imputations generated for missing values of retest results and the summary variable of ‘≥1 sex partner treated’. Clustering of outcome within clinic-period was achieved using the Blimp 2.1 software with fully conditional specification.^53^

The number of imputed datasets was proportional to the study attrition rate for the primary outcome (which was expected to have more missing values than the 2 week follow-up outcomes) – specifically the number of imputations was set to be greater than the number of percentage points for which data are missing (i.e. 55 imputations for 55% missing data). Each imputed dataset was analysed separately and the results combined using Rubin’s rules to produce a single treatment effect estimate and 95% confidence interval.

We also made use of imputed datasets to analyse the primary outcome under missing not at random (MNAR) conditions. We allowed those lost-to-follow-up to be more, and then less, likely to be chlamydia positive at 3 months than those not lost to the follow-up with the same baseline covariates. These sensitivity analyses followed the approach of Carpenter et al.^38^ to assess the robustness of our conclusions to different pre-specified assumptions about the missing data, using δ-values of loge(0.5) and loge(2). We had planned to use the same imputation datasets as for the MAR analysis for this purpose, but increased the number of imputations to 250 for the MNAR analyses because the reweighting methodology leads to estimates based mainly on only a subset of the imputed datasets.

The approach of Carpenter *et al*.^38^ is based on a logistic model in which δ represents the change in the log-odds of observing the outcome (Yi) when the response changes by one unit (here a positive in comparison to a negative test). We set R_i_ = 1 if we observe Yi and Ri = 0 if the outcome is missing. The probability of observing the outcome depends on both Yi, treatment allocation and any other baseline characteristics included in the analysis (X_i_) through the logistic model:

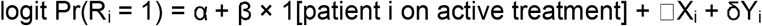

A δ value >0 implies that patients with a positive outcome are more likely to have their outcome observed while a δ<0 implies that they are less likely to have their outcome observed.

#### Exclusion of clinic-periods with suboptimal implementation

The primary outcome and first secondary outcome (≥one sex partner treated per index patient) was analysed with exclusion of clinics with very low uptake of the APT intervention. Prior to un-blinding of the statistician, the Data Manager generated a list of the % APT uptake (selection of APT by index patient as a proportion of all sex partners) for each clinic during their intervention period and a lower cut-off for adequate implementation was decided before re-analysis.

